# SEIR Filter: A Stochastic Model of Epidemics

**DOI:** 10.1101/2021.02.16.21251834

**Authors:** Martin Šmíd, Luděk Berec, Aleš Antonín Kuběna, René Levínský, Jan Trnka, Vít Tuček, Milan Zajíček

## Abstract

There are many epidemiological models at hand to cope with the present pandemic; it is, however, difficult to calibrate these models when data are noisy, partial or observed only indirectly. It is also difficult to distinguish relevant data from noise, and to distinguish the impact of individual determinants of the epidemic.

In mathematical statistics, the tools to handle all of these phenomena exist; however, they are seldom used for epidemiological models. The goal of this paper is to start filling this gap by proposing a general stochastic epidemiological model, which we call SEIR Filter.

Technically our model is a heterogeneous partially observable vector autoregression model, in which we are able to express closed form formulas for the distribution of compartments and observations, so both maximum likelihood and least square estimators are analytically tractable. We give conditions for vanishing, explosion and stationary behaviour of the epidemic and we are able to express a closed form formula for reproduction number.

Finally, we present several examples of the model’s application. We construct an estimate age-cohort model of the COVID-19 pandemic in the Czech Republic. To demonstrate the strengths of the model, we employ it to analyse and compare three vaccination scenarios.

## 1. Introduction

Whereas fundamental rationale behind mainstream epidemic models, dating back to the works of Ross [25] and Kermack and McKendrick [14] about a century ago, is relatively easy to comprehend, problems arise when we aim to parameterize such models to data for specific epidemics. The source of troubles is not only absence of clear and consistent protocols for data collection, but also factors that can never be fully intercepted, such as degrees of compliance in many adopted interventions. This is also the case for COVID-19, despite the somewhat paradoxical observation, that we now arguably have the best data on any epidemic in the history of mankind.

In regards to COVID-19, many issues arise requiring careful modelling, such as quantifying the effects of various non-pharmaceutical interventions [10, 4, 16]. Two questions are particularly interesting in this context: how effective testing and tracing has to be to counteract an intervention relaxation [19] and how stringent the interventions should be in order to compensate for partial non-compliance of the population [1]. To answer these questions, models for correctly predicting uncertainty are of vital importance.

Upon construction of realistic models, various data issues have to be coped with, starting from noise (caused both by the epidemic process itself and the data collection), insignificance (difficulty to distinguish impacts of factors from random fluctuations) or co-linearity (difficulty to distinguish two parameters with sufficient certainty). The most severe difficulty however, is that relevant data (e.g. the numbers of infectious individuals) are hidden and observed only indirectly (through the numbers of confirmed cases, for instance). Obviously, once any of these phenomena are handled insufficiently, models can provide incorrect policy recommendations.

Mathematical statistics has developed tools to handle these issues. However, to our best knowledge, there is no work systematically doing so for compartmental epidemic models. The goal of this paper is to make some progress in this area by proposing a general stochastic epidemic modelling framework.

Stochastic epidemic models do obviously exist [3]. Apart from agent-based models [2], they are commonly formulated as Markov chains [9] or stochastic differential equations [7]. However, many of these studies are theoretical, examining behavior models they construct without providing any link to real data and hence an estimation procedure. On the other hand, many other studies use elaborate filtering methods to estimate parameters of their epidemic models, but these models are largely deterministic. Their statistical models serve just as tool on the way to understand specific epidemics [24, 27]. Here, we bridge these two approaches, precisely formulating and analyzing a general stochastic epidemic model, discussing its highly practical implications, as well as providing an estimation procedure exemplified on the COVID-19 epidemic in the Czech Republic. Due to practical reasons, we develop our model as discrete in both time and state space. Although applicable to a wide class of compartment settings, we call our framework the SEIR Filter.

Models close to our model were developed by [15, 23]. To estimate their parameters, the authors used Monte Carlo simulations to evaluate likelihood functions. On contrary to these (and many other) studies, our framework enables model parameters to be estimated in a straightforward way, since the exact likelihood (or other estimating) function can be derived. This approach makes faster and more reliable parameter estimation possible. In addition to the tractability of the estimating function, our model allows for closed form formulas for expected future compartment sizes and the reproduction number. In addition, we provide simple criteria for vanishing and explosion of the epidemic, as well as for bounds that limit expected epidemic sizes given stationary imports. As we demonstrate by stylized examples, many applications of the model are possible, including derivation of implicit formulas regarding compensation measures to intervention relaxation or non-compliance, and of optimal control strategies; in this way, we touch, yet not solve, the questions mentioned above.

From the mathematical statistics perspective, we model the epidemic by a partially observed inhomogeneous heteroskedastic vector autoregression process. In line with the usual practice [8], we assume over-dispersed probability distributions both for the infections and the between-compartmental transitions; thanks to this, we are able to handle realistically not only point estimators, but also the uncertainty associated with them. Moreover, having a standard statistical output of the parameters estimation, we are able to answer questions concerning significance (via P-values^7^), possible co-linearity (via the estimator’s correlations) and the hidden compartments states (via the estimate of state-space distribution).

To demonstrate usefulness of our framework, we apply it to the actual COVID-19 epidemic in the Czech Republic. In particular, we consider four age cohorts, for each of which we model incidence, admission to – and release from hospitals and deaths. To estimate the model’s parameters, we use several partially overlapping datasets; the ability to create statistically correct estimates based on such heterogeneous datasets is, in our opinion, one of the greatest contributions of our approach. We demonstrate both in-sample and out-of-sample prediction ability of our model and, as an example of its possible use, we compare three strategies of vaccination: no vaccination, vaccination without preferences and the old-first strategy.

The paper is organized as follows. After a rigorous probabilistic formulation of the model (Section 2), we discuss its basic probabilistic properties (Section 3), its autonomous sub-models and the reproduction number (Section 4), and its asymptotic properties (Section 5). Next we introduce an age-structured version of the model (Section 6) and suggest a way to optimally control the epidemic (Section 7). Further, we discuss estimation of model parameters (Section 8). Finally, we apply our model to the COVID-19 epidemic in the Czech Republic (Section 9) and demonstrate possible model applications by comparing three possible vaccination scenarios (Section 10). Section 11 then concludes the paper.

## 2. Model Definition

Assume a population of size *s* ∈ℕ, where *s* is large. Each individual of the population is either susceptible, or finds himself in one of the compartments *S*_1_, …, *S*_*k*_. Let 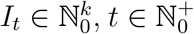, be a possibly hidden external inflow (import) of individuals into the compartments and let *Z*_*t*_ ∈ℝ^*p*^, *t* ∈ℕ_0_, be an observed exogenous process.

For any *t* ∈ℕ_0_, let 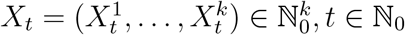, be a possibly hidden stochastic process of the compartment sizes which we define later. Let

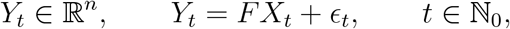

be a process of observations where *F* is a deterministic *n* × *k* matrix with rank *n*, and *ϵ*_*t*_ is a random errors vector.

Denote (ℱ_*t*_)_*t*≥0_ and (𝒢_*t*_)_*t*≥0_ the filtrations induced by (*X, Y, I, Z*) and (*Y, Z*), respectively. These filtrations may be seen as information flows, (ℱ_*t*_)_*t*≥0_ representing time evolution of all the information and (𝒢_*t*_)_*t*≥0_ representing the evolution of the observable observable. Recall that a random variable *ξ* is measurable with respect to *σ*-algebra *σ*(Ξ) generated by a random element Ξ if and only if there exists a measurable function Φ such that *ξ*= Φ(Ξ). Thus, saying that *ξ* is _*t*_-measurable (abbreviated as *ξ*_*t*_) means, that *ξ* may be devised from the values of (*Y, Z*) up to *t*.

We assume that

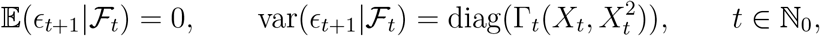

where Γ_*t*_ is a 𝒢_*t*_-measurable affine linear function (i.e. Γ_*t*_(*x, y*) = *γ*_*t*,0_ + *γ*_*t*,1_*x* + *γ*_*t*,2_*y* for some *γ*_*t*,0_ ∈ ℝ^*k*^ and *γ*_*t*,1_, *γ*_*t*,2_∈ℝ^*k*×*k*^ where all *γ*_*t*,0_, *γ*_*t*,1_ and *γ*_*t*,2_ are 𝒢_*t*_-measurable).

We define *X* recursively: We let *X*_0_ to be a possibly random vector and, for any *t* ℝℕ, we put

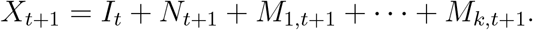

Here, 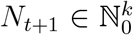 is the inflow of domestically infected individuals such that *N*_*t*+1_|ℱ_*t*_∼CPo(*A*_*t*_*X*_*t*_, *L*) where 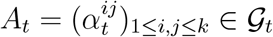 is a random matrix and, for any vector *x*, CPo(*x, L*) stands for a vector of independent Compound Poisson variables with the intensities given by *x* and the embedded distribution *L*. Observe that, by basic properties of Compound Poisson distribution,

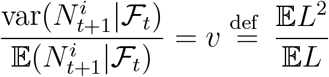

(with 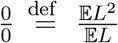), *t* ≥ 0, 1 ≤ *i* ≤ *k*. Further, for any 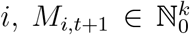 such that 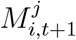 (the *j*-th component of *M*_*i,t*+1_) is the number of individuals who transited from the compartment *j* to the compartment *i* between *t* to *t* + 1. Further we assume *M*_*i,t*+1_, for any *i*, to follow Dirichlet Multinomial (DM) conditional distribution with parameters 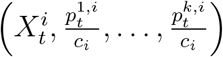 where 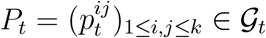 is a “mean” transition matrix and (*c*_1_, …, *c*_*k*_) are deterministic dispersion parameters.

### Remark 1.

If we assumed the individuals to change their state according to *P*_*t*_ with the transitions being conditionally independent given ℱ_*t*_, then we would get *M*_*i,t*+1_| ℱ_*t*_ as Multinomial with parameters 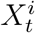 and 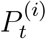 where, for any matrix Σ, Σ^(*i*)^ is its *i*-th column. In practice, however, the assumption of the conditional independence is unrealistic, as the course of infection differs between individuals and tends to cluster. The standard way of coping with this situation is using DM distribution.

The flow of individuals between states is illustrated by the following chart.

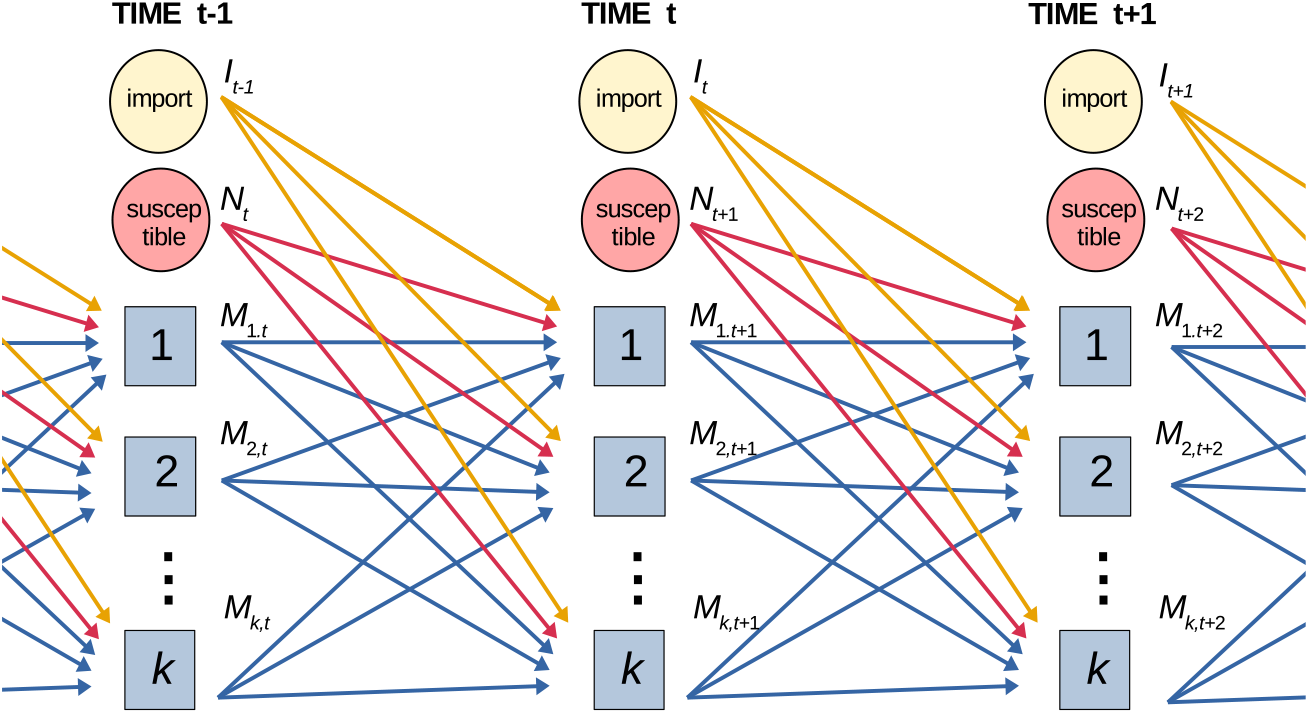

Finally, we assume *N*_*t*+1_, *M*_1,*t*+1_, …, *M*_*k,t*+1_, ∈_*t*+1_ to be mutually conditionally independent given ℱ_*t*_ (which, in words, means that all dependence between the inflows, the transitions and the observations can be explained by the state of the system at *t*). Consequently,

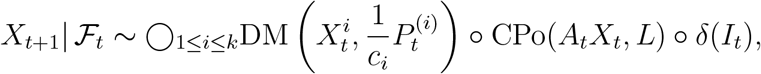

Where ◯ and ∘ stand for the summation of (mutually) independent random vectors.

### Remark 2.

The variability of the individual infectiousness may be naturally reflected by the choice of *L*. To demonstrate it, assume that only a single compartment (labeled *I*) is infectious, that all new infections fall to a single compartment (labeled *E*), and that the number of risk contacts of each infectious individual is Poisson. Let *t* ≥ 0 and denote *N*_*t*+1,*i*_ the number of the infections, caused by the *i*-th individual at *t*.

If the intensity of the contact distribution and the contagion probability were the same for all individuals, and equal to *c* and *p*, respectively, then it would be *N*_*t*+1,*i*_ ∼ Po(*cq*), *i* ∈ℕ; consequently, 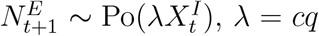, so we may put 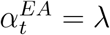 and *L* = *δ*_1_, having 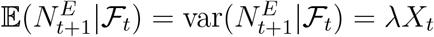.

Now consider a more realistic situation in which the infectiousness randomly varies between individuals. A standard way of modeling this situation is assuming, for each *i*, that the intensity *λ*_*i*_ of *N*_*t*+1,*i*_ is chosen from Gamma distribution, implying that *N*_*t*+1,*i*_ is negative binomial (see [28]). In particular, once *λ*_*i*_ ∼Γ(*k, θ*), and *N*_*t*+1,*i*_|*λ*_*i*_ = Po(*λ*_*i*_), we are getting that 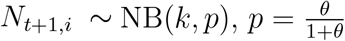, with

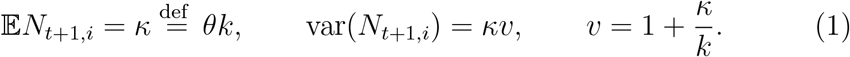

As the Negative Binomial distribution can be represented by a Compound Poisson one and as the sum of independent Compound Poisson distributions is Compound Poisson, we have that 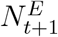 is Compound Poisson^8^ and it follows from (1) that

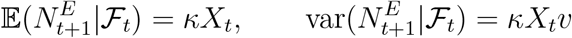

### Remark 3.

The authors of [6] claim that the total number *T* of individuals infected by a single COVID-infectious individual is *T* ∼ NB(*K, P*) where K ≐ 0.1 and *P* is such that 𝔼*T* = *R*_0_ where *R*_0_ is the basic reproduction number. As *R*_0_ of COVID-19 is generally assumed to be around 2.5 and 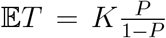, it follows that 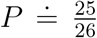. Consequently, the variance-to-mean ratio is 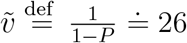. Assuming that an individual is infectious for *f* days and that his contacts are restricted by a factor *β*, we may, in light of the Compound Poisson reformulation of *T*, conclude that the number *N*_*t*+1,*i*_ of daily infected individuals is Compound Poisson with 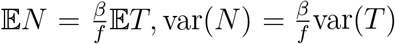, i.e. *N*_*t*+1,*i*_ and consequently 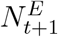 has the same variance-to-mean ratio as *T*, i.e. 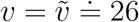.

### Remark 4.

For any *x* ∈ℕ_0_, *c* ≥ 0 and *p* ∈ [0, 1]^*k*^ such that 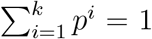, we have

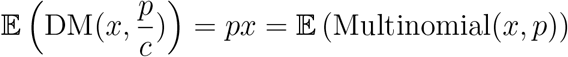

and

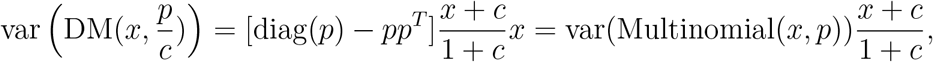

from which it is clear that 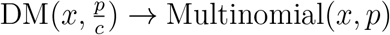 as *c* → ∞ and that the deviation of the Multinomial variance matrix grows with decreasing *c*.

## 3. Model Properties

By probability calculus, we get that

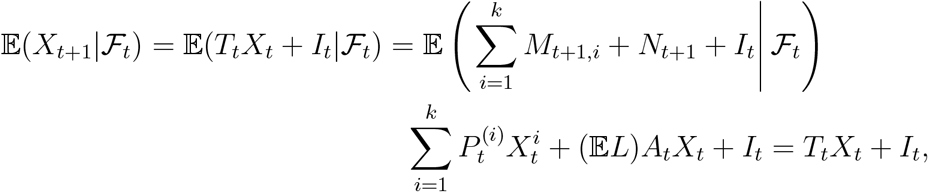

where, for any matrix Σ, Σ^(*i*)^ denotes its *i*-th column, and

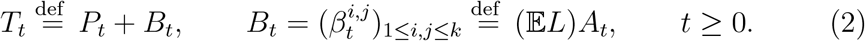

Consequently, for any *t, s* ∈ℕ_0_, *t* > *s*,

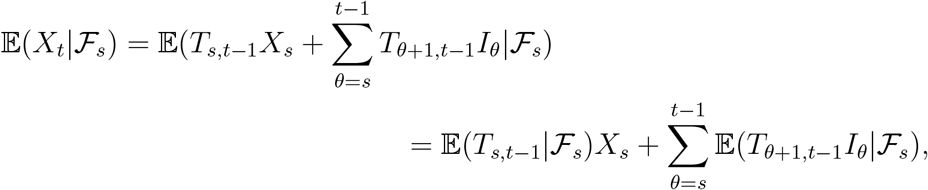

where, for any matrix process 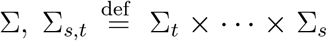 with 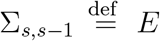 where *E* is the identity matrix.

In the special case that

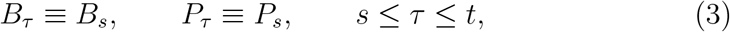

we have

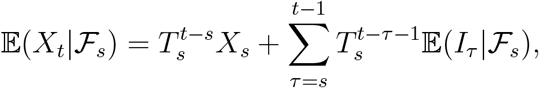

and

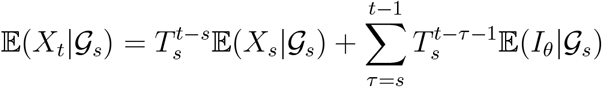

If, in addition, 𝔼 (*I*_*θ*_ *𝒢*_*s*_) *µ* for some *µ* _*s*_ and (*E − T*_*s*_) is invertible, the latter formula simplifies to

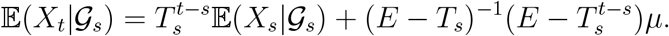

As for variance, we have

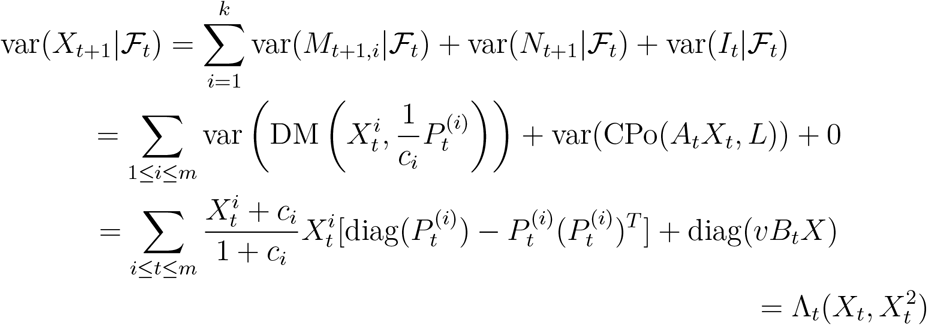

Where

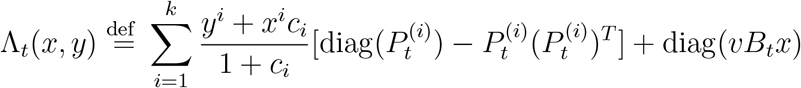

(note that Λ_*t*_ is linear in *x, y*). Consequently,

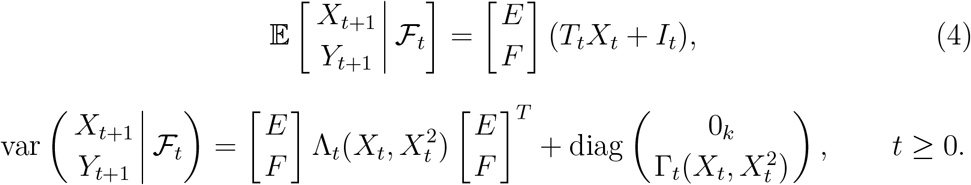

## 4. Sub-epidemics and Reproduction Number

We say that the subset of compartments *D* = *{s*_1_, …, *s*_*m*_*}* is *subepidemic* if, for any *t* and any *i* ∈ *D* and 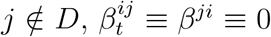 and 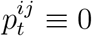. In words this means that, for any *i* ∈ *D*, the *i*-th compartment does not increase through direct infection, the infection does not depend on the compartment, and it is impossible to get to the state *i* once being outside *D*.

Let, after a possible re-ordering, *m* ∈ℕbe such that *{*1, …, *m}* is subepidemic (such *m* always exists because it can be always put to *k*). For any vector *x* ∈ℝ^*k*^, denote 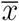 its restriction to (1, …, *m*) and, for any matrix *A*∈*ℝ*^*k*×*k*^, denote *Ā* its restriction to (1, …, *m*) × (1, …, *m*).

Observe that 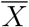 follows a slightly modified version of our model, namely

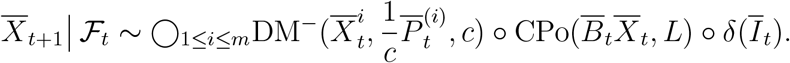

where, for any 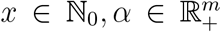 and *c* > 0, DM^−^(*x, α, c*) is the marginal distribution of the first *m* components of DM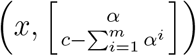 (by the aggregation property of DM).

For any *t*, we define the reproduction number *r*_*t*_ (of a subepidemic 1, …, *m*) as

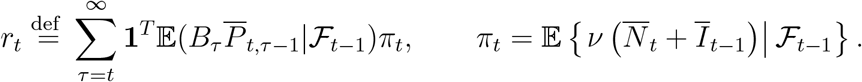

where *𝒱* is unit normalization of a vector (recall that 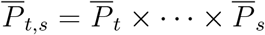). Observe that *r*_*t*_ complies with the usual definition of reproduction number as it equals to the conditional expectation (w.r.t. ℱ_*t*−1_) of the infections caused by an individual having arrived at *t*. To see it, note that *π*_*t*_ is the conditional distribution of the state in which a randomly chosen newcomer (the one brought by the import or by the infection) finds himself at *t*, and observe that, for each newcomer at *t*, the expected numbe<u>r</u> of those infected by him at *t* + 1 is given by the sum of the components of 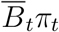, the expected number infected at *t* + 1 is given by the sum of components of 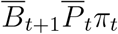 etc.

If ℱ_*t*_ = 𝒢_*t*_ (i.e. *X* is not fully observed), then the reproduction number has to be estimated, most naturally by its conditional expectation with respect to the known information:

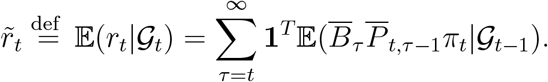

In the special case of 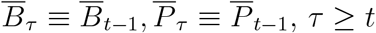, with 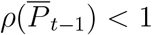 where

*ρ* is the spectral radius, the formula simplifies to

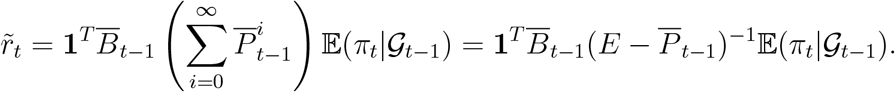

Note that, once 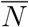 and/or *Ī* are possibly not observed, there could be difficulties computing 𝔼(*π*_*t*_|*𝒢*_*t*−1_). Although the estimate 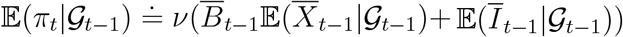 seems a straightforward choice, it is generally not unbiased due to the normalization. This problem, however, vanishes if the imports and new infections all fall into a single state (typically called exposed and labeled *E*), in which case *π*_*t*_ *≡* (1, 0, …, 0)^*T*^.

## 5. Asymptotic Behavior

Keep assuming that *{*1, …, *m}* is subepidemic. The next Proposition states conditions for vanishing, explosion and “stationary” behavior of the subepidemic.

### Proposition 5.

i. 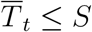 *component-wise, where S is deterministic with* 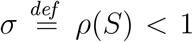 *and if 𝔼Ī*_*t*_ = *o*(*t*^−*α*^) *for some α* > 0, *then* 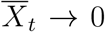 *almost surely. Here, ρ denotes the spectral radius of a matrix*.
ii. *If* 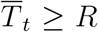 *where R is deterministic irreducible with* 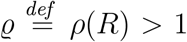 *and either* 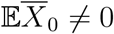 *or* 𝔼*Ī*_*τ*_ *≠ 0 for some τ, then* 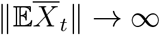.
iii. *If* 𝔼*Ī*_*t*_ ≡ *μ for some µ and* 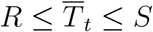 *such that* 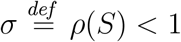, *then*

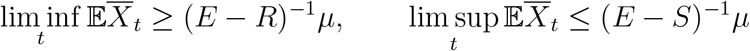

*Proof*. (i) We have

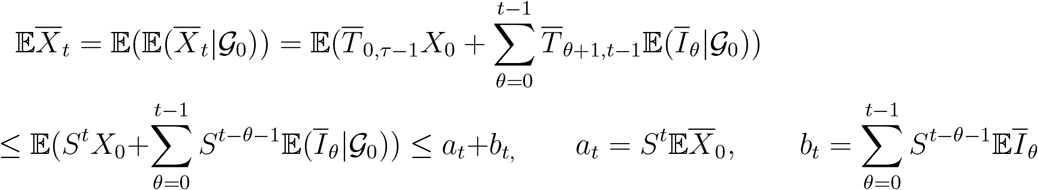

Thanks to the sub-unit spectral radius of *S*, we have *a*_*t* →_ 0. Further, by the non-negativity of *H* and the properties of convergence, there exists 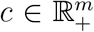 such that 𝔼*Ī*_*t*_≤*c*(*t+1*)^−*1*^. Thus, for any *ς* fulfilling *σ < ς <* 1, we get, after re-indexing the sum,

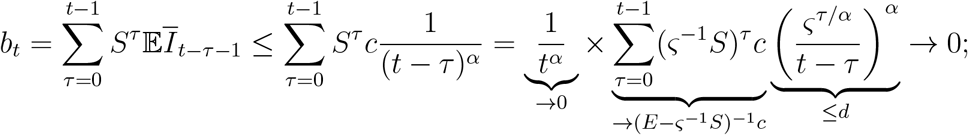

the second convergence holding because 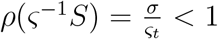, the upper bound *d* existing as 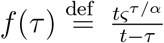 increases in *τ*= *t* − 1 and its derivative has only a single root, so we have 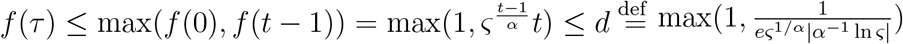 on [0, *t*−1]. Finally, thanks to the non-negativity of 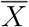, convergence of 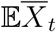 suffices for a.s. convergence of 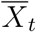.

(ii) Let 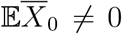 and ϱ > 1. As *R* is irreducible non-negative ϱ is its eigenvalue and the corresponding eigenvector *x* is positive by the Perron-Frobenius Theorem. Further, by the irreducibility of *T*, there exists *n* such that 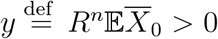 component-wise, so there exist *e* > 0 such that *y* ≥ *ex*.

Thus

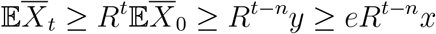

norm of which converges to infinity. The proof for 𝔼*Ī*_*τ*_ ≠ 0 is analogous. (iii)

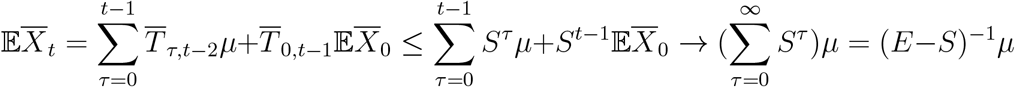

and similarly for *R*.

**Example 6**. Say there are five states *E* – exposed, *I*_*a*_ – infectious asymptomatic, who will never show symptoms, *I*_*p*_ – infectious pre-symptomatic, who will later show symptoms, *I*_*s*_ – infectious symptomatic, and *R* – removed, which includes the recovered, the dead, and the infectious isolated. We index the states by *e, a, p, s, r*. For simplicity we assume *v* = 1 which means that the new infections follow a Poisson distribution rather than a Compound Poisson one. All the infectious states are equally infectious, i.e. 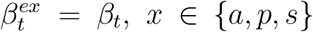, where *β* is a *𝒢*_*t*_-adapted process. The probability that the exposed transits to *{a, p}* is *σ*, the probability of completely asymptomatic course is *α*, the probability of transition from *I*_*p*_ to *I*_*s*_ is *ς*. Further, the probability of ending *I*_*a*_ or *I*_*s*_, by natural causes (recovery, end of infectiousness, death in case of *I*_*s*_) is ϱ _*a*_, ϱ_*s*_, respectively. Finally, the probability that a symptomatic individual isolates himself is *η* and the probability that the individual finding herself in state *E, I*_*a*_, *I*_*p*_ or *I*_*s*_ is isolated is *θ*_*t*_ for some 𝒢_*t*_-adapted process *θ*^*i*^. The situation is illustrated on the following Figure:

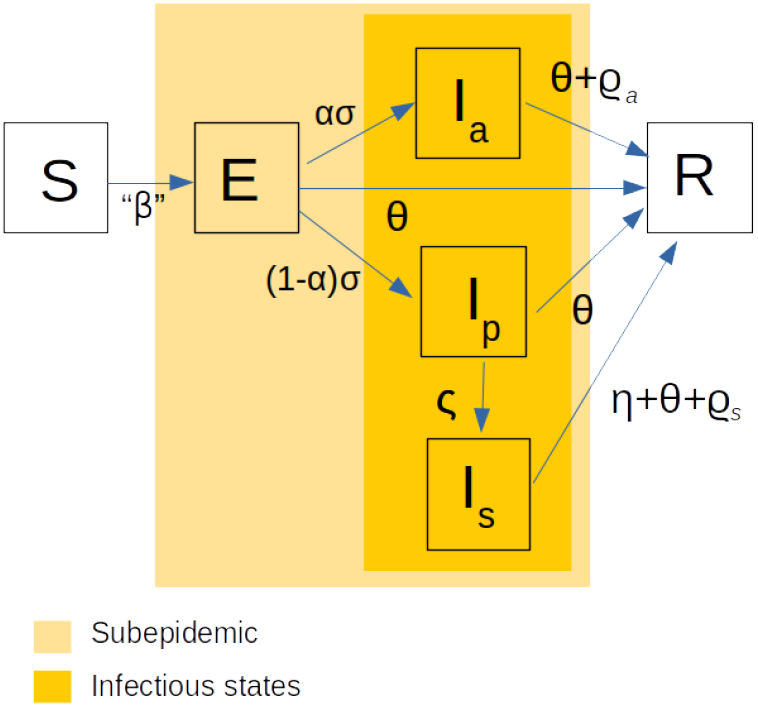

If we neglect (small) joint probabilities of natural exits from the infectious states and the isolations, we get

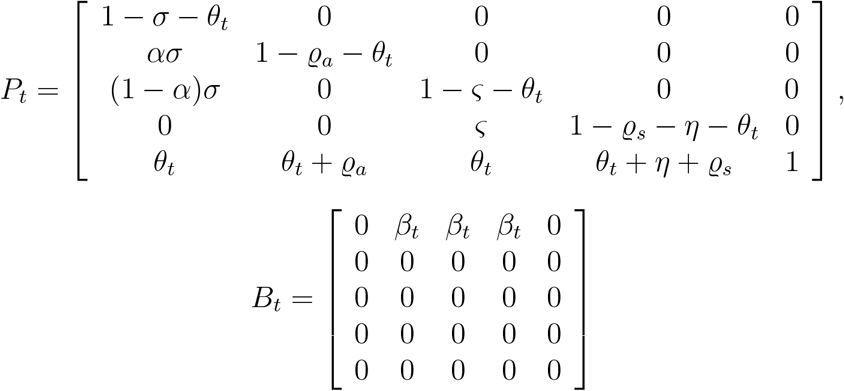

Clearly, we can put *m* = 4 (the first four states form a sub-epidemic), getting

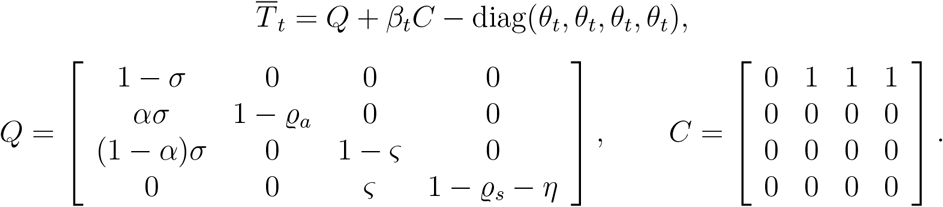

By the well known rule, 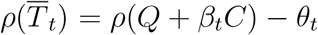. We consider two ways of decreasing the spectral radius: decreasing the infection rate *β*_*t*_ (typically by wide counter-epidemic measures) and increasing the isolation rate *θ*_*t*_ (e.g. by strengthening the testing and tracing capacity).

Once there is a “target” spectral radius *ρ*_0_, all the combinations of *β* and *θ* yielding 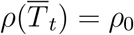 fulfill *ρ*_0_ + *θ*−*ρ*(*Q* + *βC*) = 0, which gives a “marginal rate of substitiution” 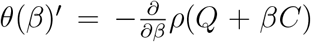 of the infectiousness by the isolation, i.e. how much we have to increase the isolation speed when we release the restrictions.

**Example 7**. Assume that a fraction *ν* of the population is non-compliant, which means that, once a restriction on social contacts is imposed, they obey it only partially. Assume that, without restrictions, the population is mixed which means that each individual, compliant or not, has, up to a constant, (1−*ν*) contacts with the compliant individuals and *ν* contacts with the noncompliant ones. Once there is a measure imposed, the compliant individuals restrict their opportunities to contacts by *ϕ*, while the non-compliant ones do so only to *f* (*ϕ*) > *ϕ*. As a result, the compliant ones will have, up to a constant, *ϕ*^2^(1−*ν*) contacts with the compliant ones, *ϕf*(*ϕ*)*ν* contacts with the non-compliant ones, while the non-compliant will have *ϕf* (*ϕ*)(1−*ν*) and *f* (*ϕ*)^2^*ν* contacts with the compliant, non-compliant, respectively.

Assuming a simple epidemic model with compartments *I*_*c*_ - infected compliant, *I*_*n*_ - infected non-compliant, and *R* - removed, with the course of infection being the same for both the compartments such that 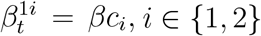, where *β* is a constant and *c*_*i*_ is the number of contacts of the *i*-th sub-population, this gives

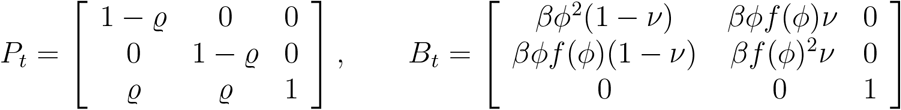

where ϱ is a removal rate (perhaps consisting of an artificial and a natural part). This gives

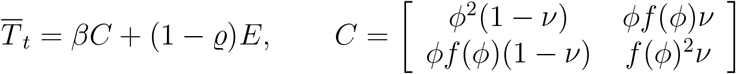

with

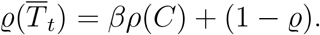

As the characteristic polynomial of *C* is

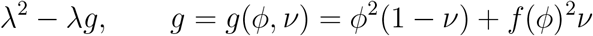

we clearly have *ρ*(*C*) = *g*.

Now say that our goal is to decrease 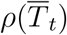 to a predetermined value *r* by finding appropriate *ϕ* = *ϕ* (*ν*). In order to do so, we have to solve

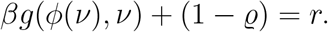

Clearly, 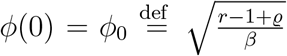. For *ν* > 0 we get, by the Implicit function theorem,

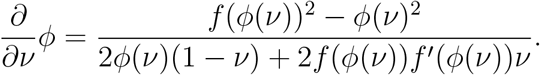

Note that the derivative depends neither on *r* nor on ϱ. Thus we can easily compute how the non-compliance influences strictness of the necessary restrictions. For instance, by the first-order Taylor expansion at *ν* = 0, we get

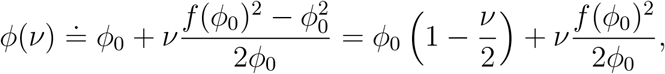

roughly holding for *ν* close to zero.

## 6. Cohort Model

In the present Section, we assume the population is split into *r* (age) cohorts of sizes *s*_1_, …, *s*_*r*_, *s*_1_ +…+ *s*_*r*_ = *s*. The members of each cohort may be either susceptible or belong to one of *κ* analogous compartments. Naturally assuming that individuals do not migrate between cohorts, we get the overall transition matrix as

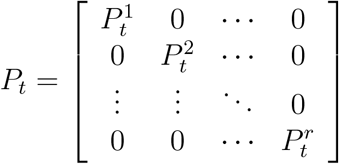

where 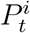 are *κ* × *κ* cohort transition matrices, 1 ≤ *i* ≤ *κ, t* ≥ 0. Note that once there are dispersion parameters 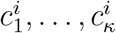 associated with each matrix 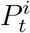 (meaning that, once the *j*-th compartment of the *i*-th cohort size *x*, the transfers from the cohort to the cohort’s compartments follow 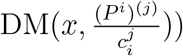 the dispersion parameters of the overall model are 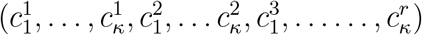.

We assume that the contagions can happen across cohorts and that the probability of contagion, i.e. the transfer of a susceptible individual to the *i*-th compartment of the *p*-th cohort, upon a risk contact with a member of the *j*-th compartment of the *q*-th cohort does not depend on *p* or *q*, being equal to 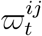. Further we assume that, on average, a member of the *p*-th cohort has *ν*^*pq*^ risk contacts with the *q*-th cohort, assuming that the number of contacts with the infectious compartments (those with non-zero 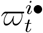) is equal. Under these assumptions, the probability of a transfer of a susceptible individual from cohort *p* into the *i*-th compartment is roughly 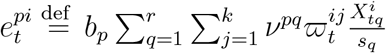, where 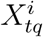 is the size of the *i*-th compartment of cohort *q* and *b* is a constant. Consequently, the total number of the infections in the *i*-th compartment of the *p*-th cohort will be 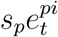 which gives

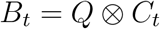

where

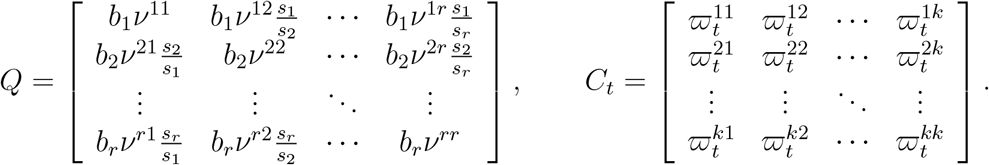

It may be convenient to re-parametrize the model by multiplying *C* by a constant and adjusting constants *b*; in this case, however, the components of *C* cease to be interpretable as probabilities.

## 7. Optimal Control of the Epidemic

Assume the settings of Example 6 and assume that *X*_0_ is known. Our aim is to minimize the size of the epidemic at time *t* given that we are ready to pay a given price *c*_0_. We assume that, to achieve infection rate *β*, a cost *γ*(*β*) has to be paid where *γ* is a strictly decreasing convex positive function defined on (0, *β*_0_] with *γ* (*β*_0_) = 0 and *γ* (0−) = ∞. Further, to achieve the isolation rate 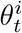 in the *i*-th compartment, the price 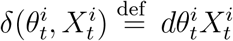 has to be paid where *d* is a constant. This reflects the real-life situation in which the cost of global restrictions does not depend on the infection size while the cost of isolation does, for instance through the number of call-center workers involved in tracing.

Our problem is to find

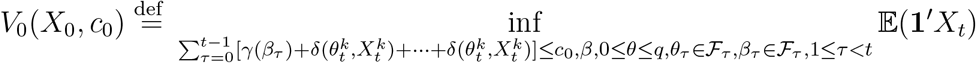

where *q* is the diagonal of *Q*. The problem may be rewritten by means of Bellman equations

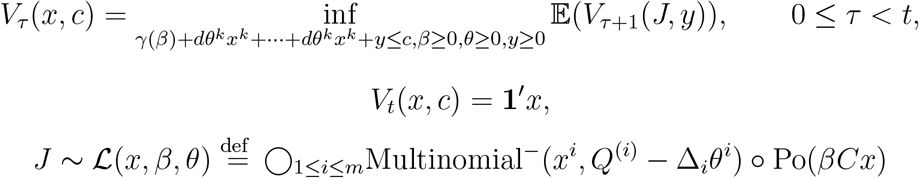

where Δ_*i*_ is the vector with the unit component on the *i*-th place and zeros otherwise.

Though the final problem is convex,

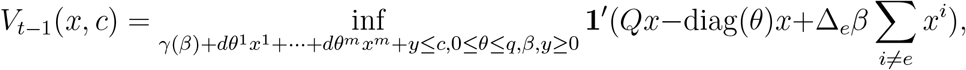

its objective function is not jointly convex in (*u, v, x, c*), so the convexity of *V*_*t*−1_ is not guaranteed. Moreover, as neither the optimal solution nor the expectation of the objective functions of all but the last problem are analytically tractable, it is necessary to resort to approximations. To this end, we can use the sample mean approximation

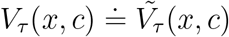

where 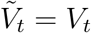 and, recursively,

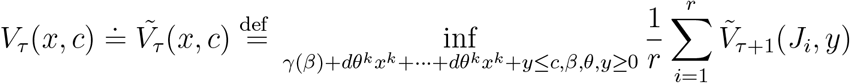

where *J*_1_, …, *J*_*r*_ is an i.i.d. sample from *ℒ*(*x, β, θ*).

## 8. Estimation

For any stochastic process *A* and integers *s*≥*t*, denote Â _*s*_ | *t=𝔼(A*_*s*_ | *𝒢* _*t*_.Let *s*>*t*. When *T*_*τ*_∈*𝒢*_*t*_, *t < τ*≤*s* −1 (which is trivially true if *s* = *t* + 1), we get that

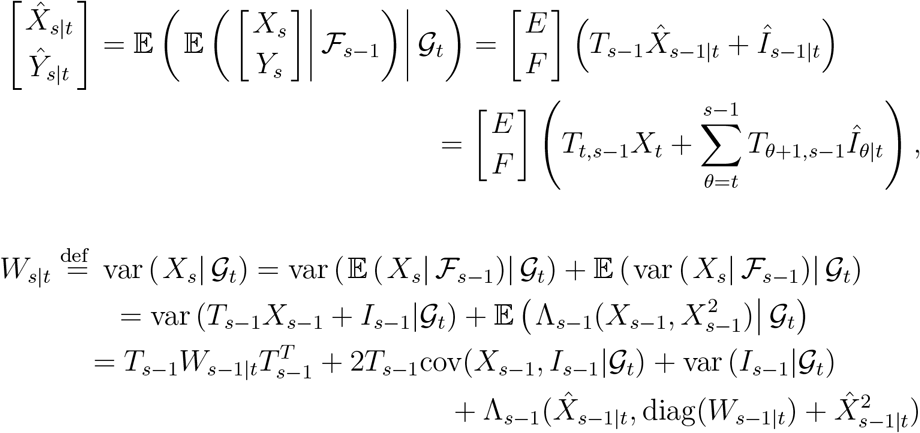

(we have used linearity of Λ_*s*−1_, thanks to which 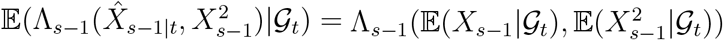 and the well known formula var(*X*) = 𝔼*X*^2^ −(𝔼*X*)^2^). Consequently,

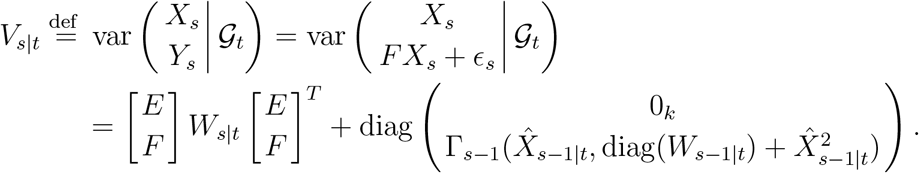

Unfortunately, due to the non-Gaussianity, we have analytical formulas for none of *X*_*t*|*t*_, *I*_*t*|*t*_ and *W*_*t*|*t*_, so we can formulate neither the likelihood function nor a least square estimate. From the computational point of view, two equivalent ways to cope with this are using estimates of the conditional expectation and variance, or normally approximating the residuals. We choose the latter way here and assume that 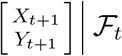 is normal, with mean given by (4) and

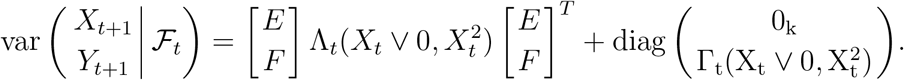

Moreover, we assume that *I*_*t*_ ∈*𝒢*_*t*_, *t* ≥ 0 (i.e. the import is observable). Given these assumptions, we have, by the well-known formula (see e.g. [5]),

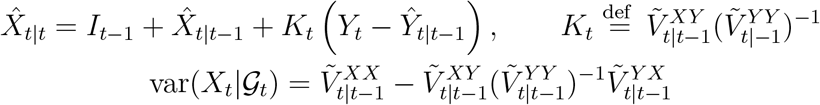

where 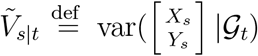 given the normal approximation. Note that *K*_*t*_ may be seen as a conditional version of the Kalman gain matrix.

If 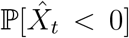 is negligible (which is typically true when modeling large epidemics), then we can neglect truncation in the formula for the variance approximate 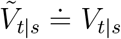. This further gives

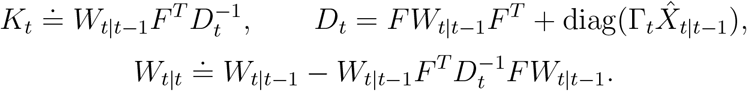

Assume that *F* = *F* (Θ_0_), *P*_*t*_ = *P*_*t*_(Θ_0_), *B*_*t*_ = *B*_*t*_(Θ_0_), Γ_*t*_ = Γ_*t*_(Θ_0_), *I*_*t*_ = *I*_*t*_(Θ_0_), and *c* = *c*(Θ_0_), where Θ_0_ ∈ℝ^*r*^ is an unknown parameter. For its estimation, it is possible to use either nonlinear least squares, i.e.

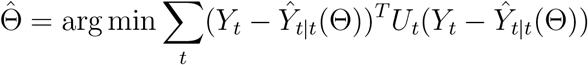

where *U*_*t*_ ∈*𝒢*_−1_ is a suitable weighting matrix, or

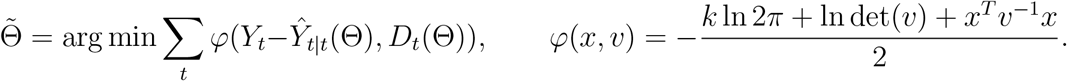

Both these estimators are consistent and asymptotically normal under some conditions, see [12] and [13], respectively. Verifying these conditions for our model is, however, beyond the scope of this introductory study and remains the topic of a future research.

It should be noted that our proof of Proposition 5 is not valid for the approximate model, as 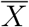 is not necessarily positive given the Normal approximation.

## 9. Application to The COVID Pandemics in the Czech Republic

We applied our model to the data from the Czech Republic epidemic between February 2020 and January 2021. We considered a widely generalized version of the model from Example 6, compartments of which are shown in Figure 2.

**Figure 1:**
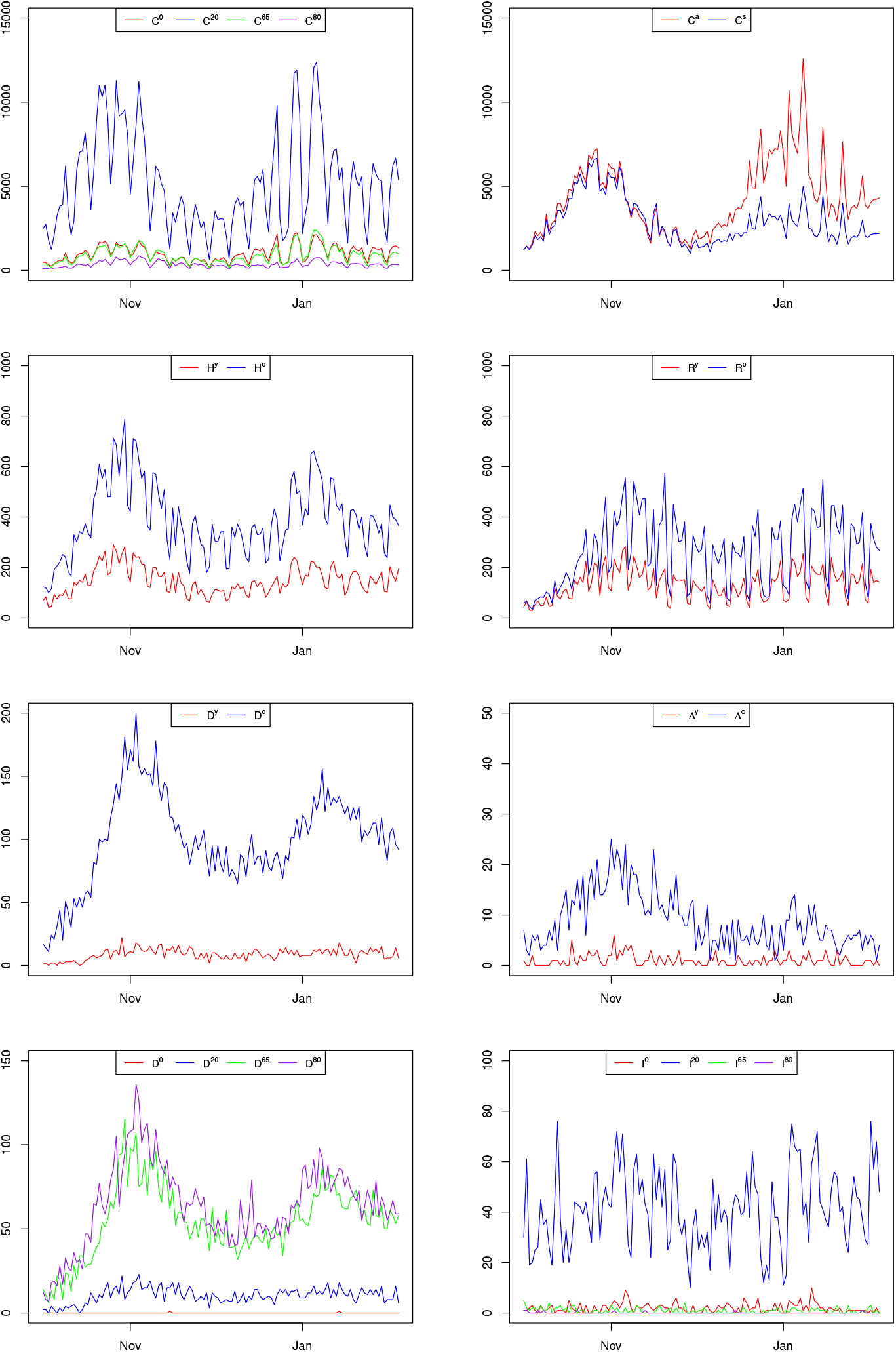
Inputs of the Czech COVID-19 model

**Figure 2:**
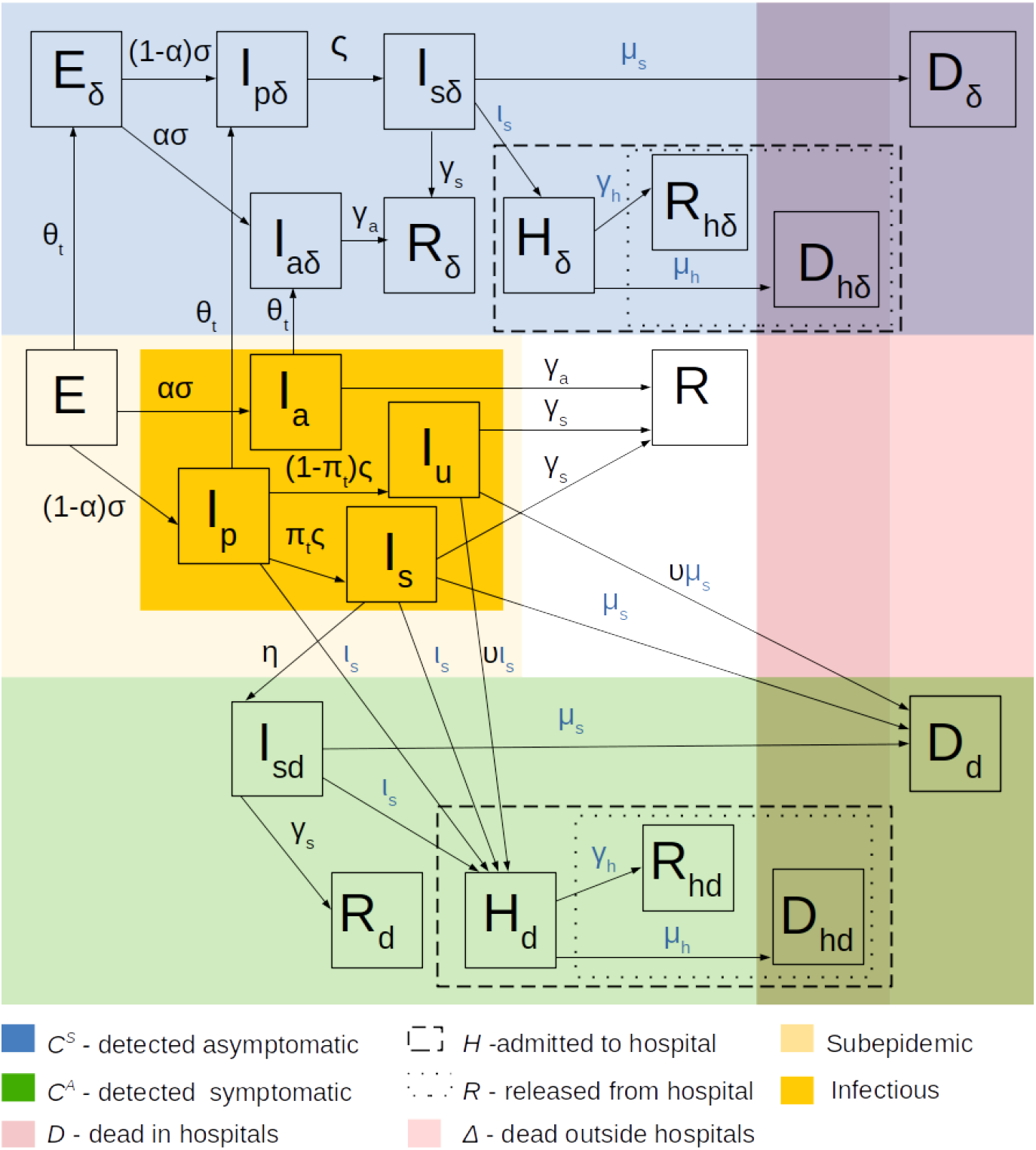
Structure of the Czech COVID-19 model.

To reflect the fact that many cases are (intentionally) undetected, we added the compartment *I*_*u*_. Members of *I*_*u*_ can be detected only if they are hospitalized or die, otherwise they never undergo testing. Further, to all the compartments from Example 6, we added their “detected” versions, distinguishing detection when being asymptomatic (subscript *δ*) or symptomatic (subscript *d*). We distinguish three “removed” states: recovered (*R*), hospitalized (*H*) and dead (*D*). Assuming that each individual who gets to the hospital is detected upon entry, we have two versions of *H* states: detected when symptomatic (*H*_*d*_) and detected when asymptomatic (*H*_*δ*_). Similarly, we assume that each dead is detected and we distinguish whether the individual dies in- or outside of hospital; consequently, we have four “dead” states: *D*_*hd*_, *D*_*d*_,*D*_*h δ*_ and *D*_*δ*_. The case of *R* is analogous with the exception that recovery can happen without detection, so we have five versions: *R, R*_*dh*_, *R*_*d*_,*R*_*δh*_, and *R*_*δ*_.

We distinguish four age cohorts: 0 to 19 years, 20 to 64 years, 65 to 79 years and more than 80 years; consequently, having four compartments for each state, we have 4×21 = 84 compartments in total.

Some transition parameters (components of *P*_*t*_) are common for all compartments (black symbols in Figure 2), the others are specific for each compartment (blue symbols). Some parameters are constant in time, some (these with index *t*) are time varying. The values of some parameters were externally determined, some were estimated. The detailed description of parameters, way of their determination, computation or estimation, and their estimated values can be found in Appendix, Table 2 and below. Here we only mention that we take

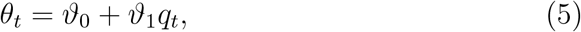

where *q*_*t*_ is the observed probability that an infectious individual is accepted to hospital without being previously detected, which, by the Bayes rule, is, up to a constant, equal to the overall detection probability. The rate *π*_*t*_ is set so that the detection probability holds in the model (see the Appendix). The value of *q*_*t*_ is publicly available as it is, as of February 2021, one of the input of the national counter-epidemic system PES.

**Table 1:** Outcomes of Vaccination Scenarios

|  | novac | middle | elderly | novac<br>-middle | novac<br>-elderly | middle<br>-elderly |
| --- | --- | --- | --- | --- | --- | --- |
| Total incidence | 872050 | 814270 | 843350 | 57780 | 28700 | -29080 |
| Total deaths | 15712 | 12413 | 11710 | 3298 | 4001 | 703 |
| Average hospitalization | 7067 | 5906 | 5753 | 1160 | 1314 | 154 |

**Table 2:**
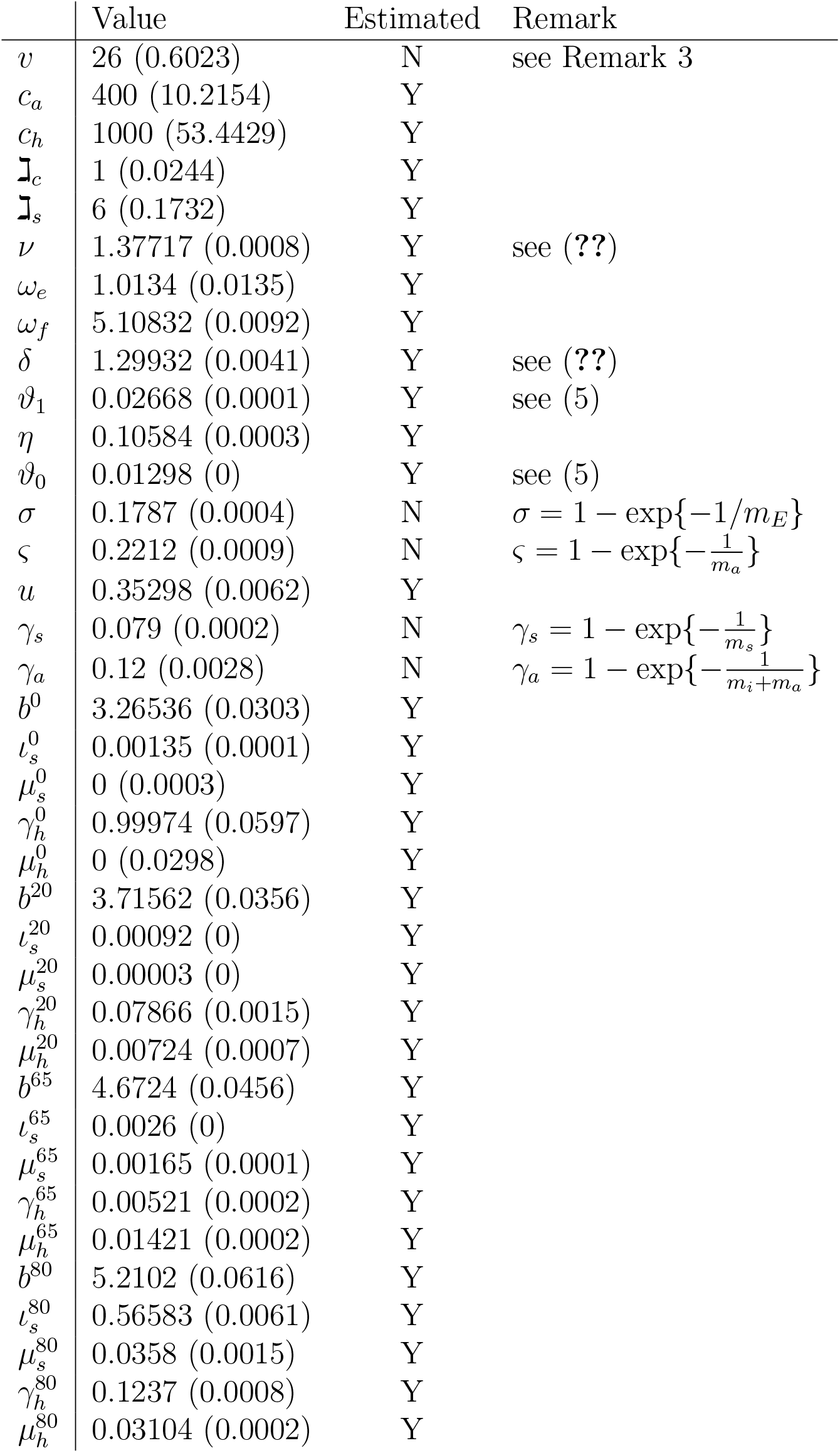
Parameters of the Czech COVID Model

The normalized “mixing matrix” *Q* has been computed from the estimate of the Czech overall contact matrix by [22], and is listed in Appendix, Table 3. In line with [21], reporting 3 −25 times lower secondary attack rate of asymptomatic individuals, we assume that the probability of infection by *I*_*a*_ is four-times less in comparison with *I*_*p*_, *I*_*s*_ and *I*_*u*_. Further, we assume the dependence of infectiousness on the contact restriction and personal protection, and we reflect the natural immunization. Namely, we have

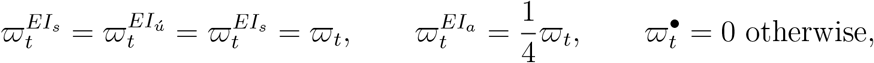

where

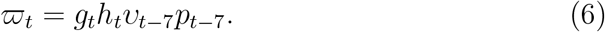

**Table 3:** Popluation of Cohorts and Mixing Matrix *Q*

| Cohort | 0 | 20 | 65 | 80 |
| --- | --- | --- | --- | --- |
| Population | 2188232 | 6374077 | 1690530 | 441100 |
| $Q$ | 0 | 20 | 65 | 80 |
| 0 | $b^0 \times 0.54$ | $b^0 \times 0.11$ | $b^0 \times 0.02$ | $b^0 \times 0.02$ |
| 0 | $b^0 \times 0.6$ | $b^0 \times 0.92$ | $b^0 \times 0.12$ | $b^0 \times 0.12$ |
| 65 | $b^{65} \times 0.09$ | $b^{65} \times 0.1$ | $b^{65} \times 0.14$ | $b^{65} \times 0.14$ |
| 80 | $b^{80} \times 0.02$ | $b^{80} \times 0.02$ | $b^{80} \times 0.02$ | $b^{80} \times 0.06$ |

Here, *𝓋*_*t*_ is the average number of weekly risk contacts of an individual, *p*_*t*_ is the reduction caused by personal protection, ht 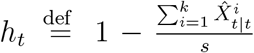 is the adjustment for immunization (we assume that once infected individuals cannot be infected again), and *g*_*t*_ is the adjustment for new mutations, see Appendix for details.

The values of *𝓋*_*t*_ and *p*_*t*_ were taken from [20] which is a longitudinal study, inquiring a panel of 3000 respondents about their (weekly) risk contacts, observance of several personal protection measures (see Appendix), and some other variables. While the value of *𝓋*_*t*_ is being directly questioned by the survey, we compute the level *p*_*t*_ of personal protection as

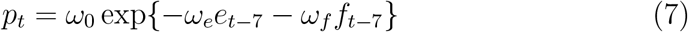

where *e*_*t*_ is a certain linear combination of observation rates of various personal protective measures, *f*_*t*_ is the reported level of fear caused by the pandemic and *ω*_0_, *ω*_*e*_ and *ω*_*f*_ are constants (see Appendix for further explanation).

For better comparison with other models, we use normalized version of the personal contacts level 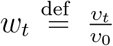 where *𝓋*_0_ is the pre-pandemic value of *𝓋*, so we have, up to constant multiplication,

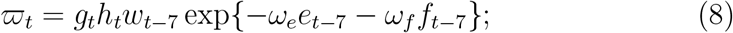

with the corresponding constant becoming part of estimated parameters *b*_0_, *b*_20_, *b*_65_ and *b*_80_.

As for the dispersion parameters, we assume *v* = 26 (see Remark 3) and, for each cohort *i*, we put 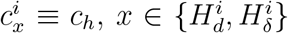 (all “hospital” states) and 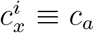 otherwise. This restriction of the parameter space was done in order to avoid over-parametrization, but to reflect the larger variability of incidences and hospitalizations in comparison with the m Ĩ ortality data (note that dead mostly come from hospitals). Note also that, for terminal states, the dispersion parameters have no effect, as their “target” distribution is Dirac.

We used the following daily data series as inputs (see Figure 1):

*Ĩ*^0^,*Ĩ*^20^,*Ĩ*^65^,*Ĩ* ^80^ - numbers of positively tested with infection abroad of individual cohorts, serving as imports (see below) and the following series which we consider as observations (variables *Y*) in our model:

*C*^*A*^, *C*^*S*^ *-* incidences of asymptomatic and symptomatic

*C*^0^, *C*^20^, *C*^65^, *C*^80^ - incidences in individual cohorts

*D*^0^, *D*^20^, *D*^65^, *D*^80^ - deaths in individual cohorts

*H*^*y*^, *H*^*o*^ *-* admissions to hospitals of individuals from the first two cohorts and second two cohorts

*R*^*y*^, *R*^*o*^ *-* numbers of released from hospitals of the first and second two cohorts

*D*^*y*^, *D*^*o*^ *-* dead in hospitals of the first and second two cohorts

Δ^*y*^, Δ^*o*^ *-* dead outside hospitals of the first and second two cohorts

The data come from the public repository of the Ministry of Health of the Czech Republic and from the The Institute of Health Information and Statistics of the Czech Republic, where the former is publicly available while the latter is available only to research institutions. See Figure 1 for overview.

The data had to be pre-processed. First of all, series *C, H* and *R*, showing strong weekly pattern, have been de-seasoned. Second, as the total reported incidence exceeds (by about 2 percent) the personal level data (probably because details are not always known), we took the former as decisive. Next, as the reports of symptomatic an asymptomatic results of tests give slightly less totals then the incidence, we took only ratios from the former multiplied by the total incidences as *C*^*a*^ and *C*^*s*^. Finally, as the total reported number of actually hospitalized, which we denote by *L*, exceeds (by units of percents) the value *H* − *R* coming from the anonymized hospitalization data, we adjusted series *H* and *R* so that *L* ≐ *H* − *R* (doing it exactly is impossible, as plain multiplication could cause unrealistic values of *H* and *R*).

Only three series take part as exogenous variables (values *Z*) in our model:

*w* - normalized contact restriction

*e* - personal protection index

*f* - level of reported fear

*q* - probability of detection in hospital

(see explanations above).

The matrix *F*, transferring compartment sizes *X* to observations *Y*, is zero-one. Which components are equal to one can be partially devised from Figure 2, the exact definition of *F* is given in Appendix, Figure 6. The lack of a row of *F* corresponding to *C*^*a*^ is intentional as *C*^*a*^ is redundant (equal to *C*^0^ + *C*^20^ + *C*^65^ + *C*^80^ − *C*^*S*^) and would cause linear dependence in *F* if included.

Assuming the imports only to the state *E*, we take,

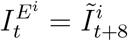

for each cohort *i* (the time-shift reflects the delay in reporting). Reflecting the notorious unreliability of incidence data, we put

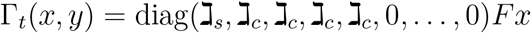

where ℷ_*s*_ and ℷ _*c*_ are estimated parameters (note that the non-zero rows of Γ_*t*_ correspond to *C*^*S*^, *C*^0^, *C*^20^, *C*^65^, *C*^80^ respectively).

For estimation, we used Weighted Least Squares applied to the seven day forecasts with weekly average increments as weights:

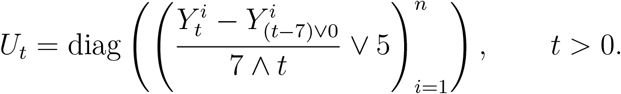

Due to the possibly different nature of the first wave (until May) and irregular behavior of the pandemic in summer (caused by small incidence with local outbursts), we used only data from October onward for the (final) estimation. To get “reasonable” initial state for our estimate, we roughly estimated the model for the whole period of the pandemic and took its state from the end of August as the initial state for the final estimation; to minimize the influence of the initial value, however, the WLS criterion was computed only for the period from October 1, 2020 till January 21, 2121.

Unfortunately, the estimation did not directly lead to quality results. The reason was that minimization with free *c*_*u*_, *c*_*d*_, *γ*_*s*_ and *γ*_*c*_ (dispersion) parameters tends to underestimate forecast variances, which leads to unrealistically narrow confidence bounds. In order to get more realistic variances, we adjusted the risk parameters of the estimate so that the variance of standardized (one day forecast) residuals were close to one; we use grid search here. Then we re-estimated the model by WLS with the adjusted risk parameters fixed. Yet the value of the new WLS minimum is about 25 percent higher than the original one, the variances in the new model are realistic. Finding a fully automatic estimation procedure getting realistic variances, perhaps by regularization with respect to variance of residuals, is a challenging topic of future research. The C++ code for the WLS estimation, employing NLopt library for minimization, is available on https://github.com/cyberklezmer/seir/tree/ejor.

The standardized residuals (i.e. the residuals divided by the standard error of the estimate) of the seven-day predictions for the individual observables, may be seen in Figure 3. Figure 4, on the other hand, shows the predictions fit of the total incidence 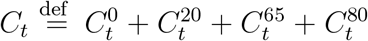, the total number of deaths 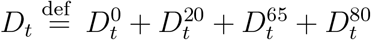 and the actual occupancy of hospitals 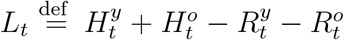. For each variable, the left-hand graph shows the fit of the (in-sample) seven day predictions, the right-hand graph depicts the (out of sample) predictions for 14 days following Jan 21.

**Figure 3:**
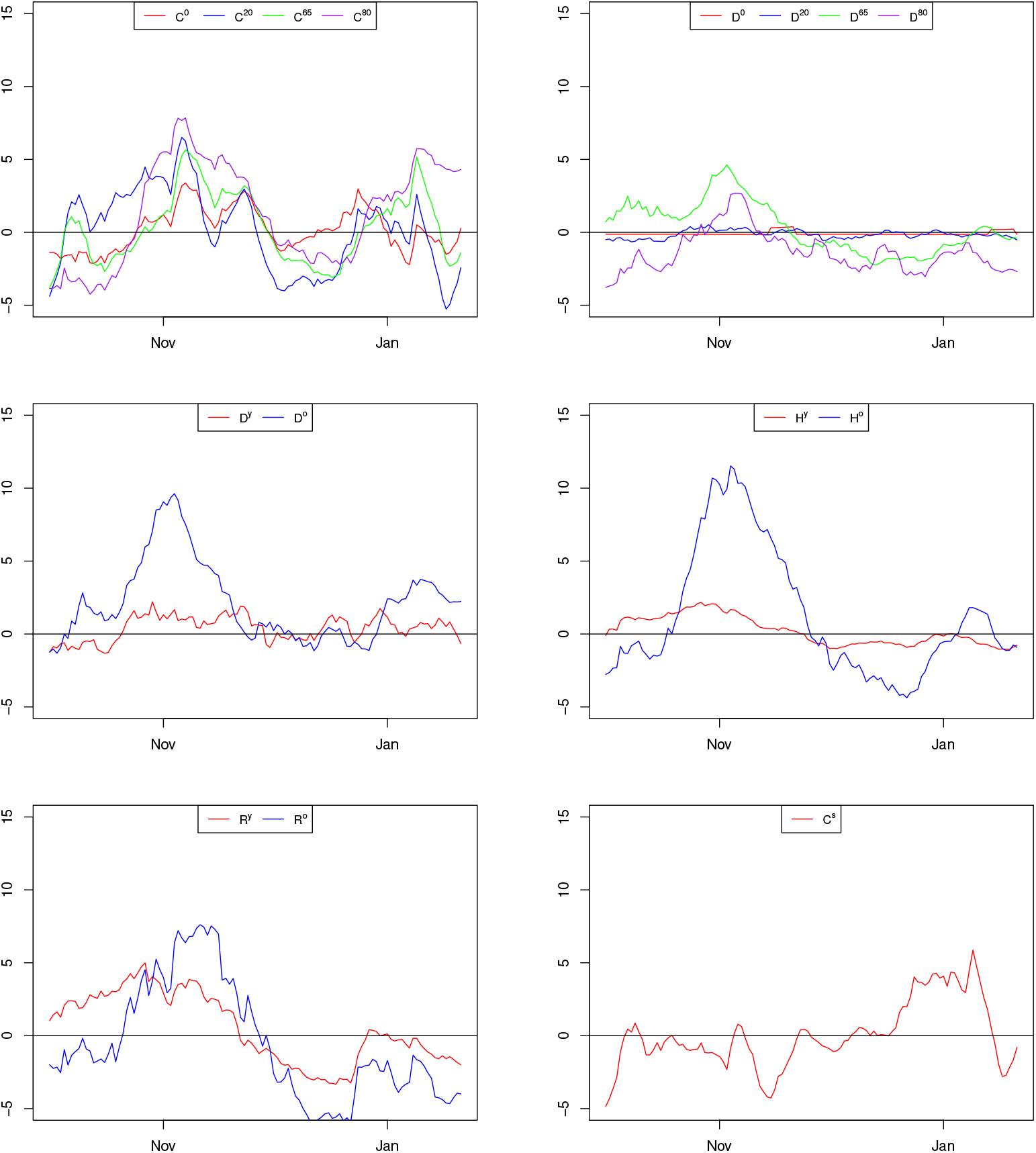
Residuals of the Czech COVID-19 model.

**Figure 4:**
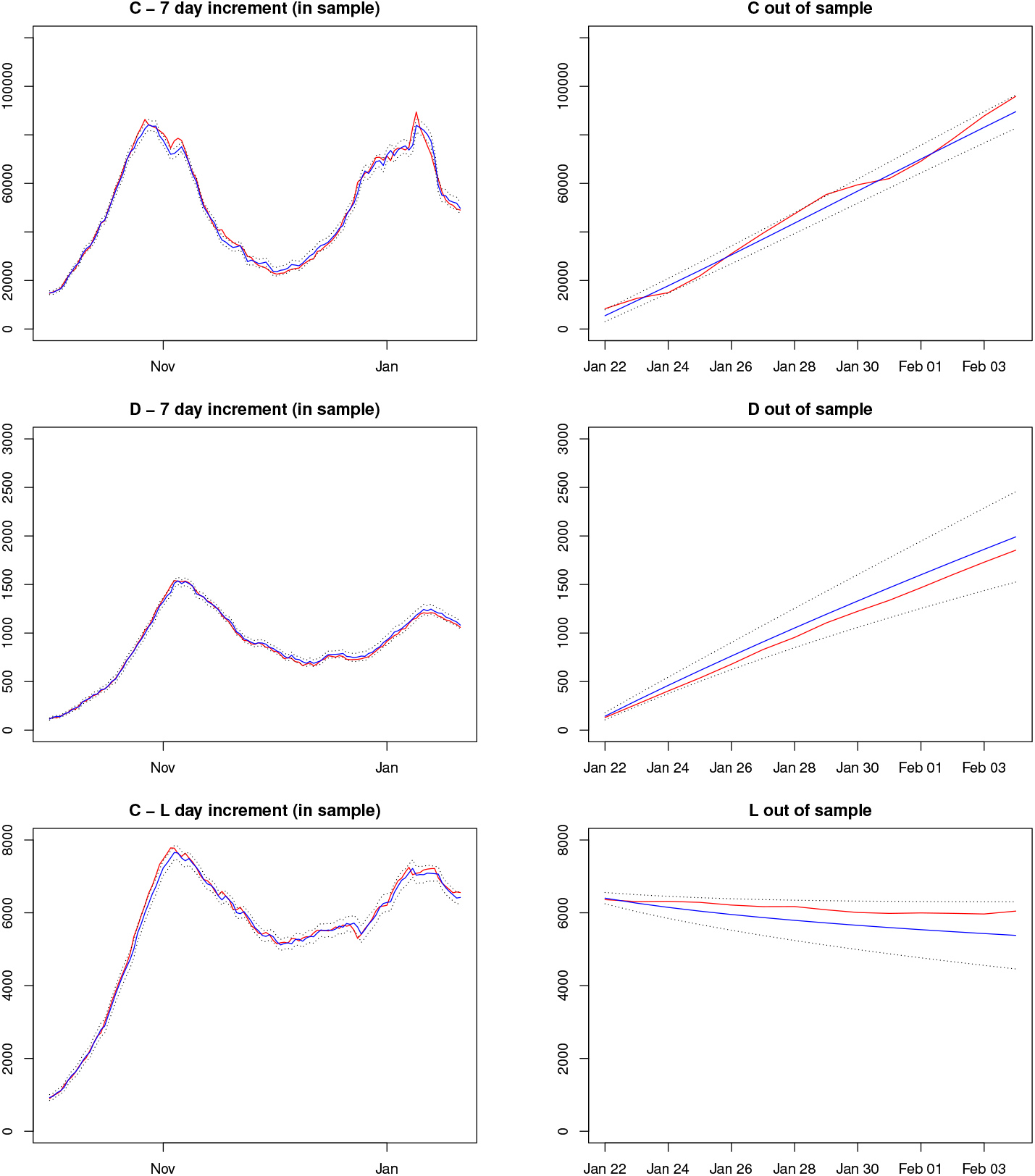
Predictions of the Czech COVID-19 model. Red – actual, Blue – predicted, dotted – 95 % confidential bound.

## 10. Vaccination Experiment

Finally, as a demonstration of applicability of our model, we evaluated outcomes of three vaccination scenarios

### novac

The infectiousness *ϖ*_*t*_ increases by 50% on January 22 (by emergence of a new mutation and/or release of the counter-epidemic measures) and keeps being constant until the end of May

### elderly

The infectiousness rises as above, but 30000 shots of vaccine a day will start to be applied on February 1st. Vaccines are distributed to the cohort 80+ first and, as late as the whole cohort is vaccinated, vaccines start to be given to the cohort 65-79. After 35 days, each vaccinated individual gets a second shot (these shots do not count to the daily number 30000).

### middle

The situation is the same as in “elderly” scenario with the difference that only 15000 vaccines a day are given to the oldest, the rest are given to cohort 20-64 (perhaps infrastructural workers).

As for the vaccine effect, we (conservatively) assume that vaccinated individuals can be infected the same way as if they were not vaccinated; however, the probability of the asymptomatic course rises: 14 days after the first shot to 0.9 for the first two cohorts (0-64), or to 0.7 for those who are older than 65. After 14 days from the second shot, the probability further rises to 0.95 and 0.8, respectively. We insert these effects into the model by adjusting the overall probability *α* of asymptomatic course in individual cohorts according to the number of vaccinated therein.

The results of simulation are shown in Figure 5 and Table 1. Not surprisingly, vaccination saves all the infections, deaths and hospital beds. Prioritizing the older saves lives and beds, however, adds infections (which partially due to greater efficiency of the vaccine for the younger cohorts, maybe also due to greater number of contacts of the young). The most important message, however, is that vaccination is not a “silver bullet” solving everything – there are still large numbers of dead even with vaccination, which could be saved if the infectiousness was reduced, too, perhaps by some unpopular measures.

**Figure 5:**
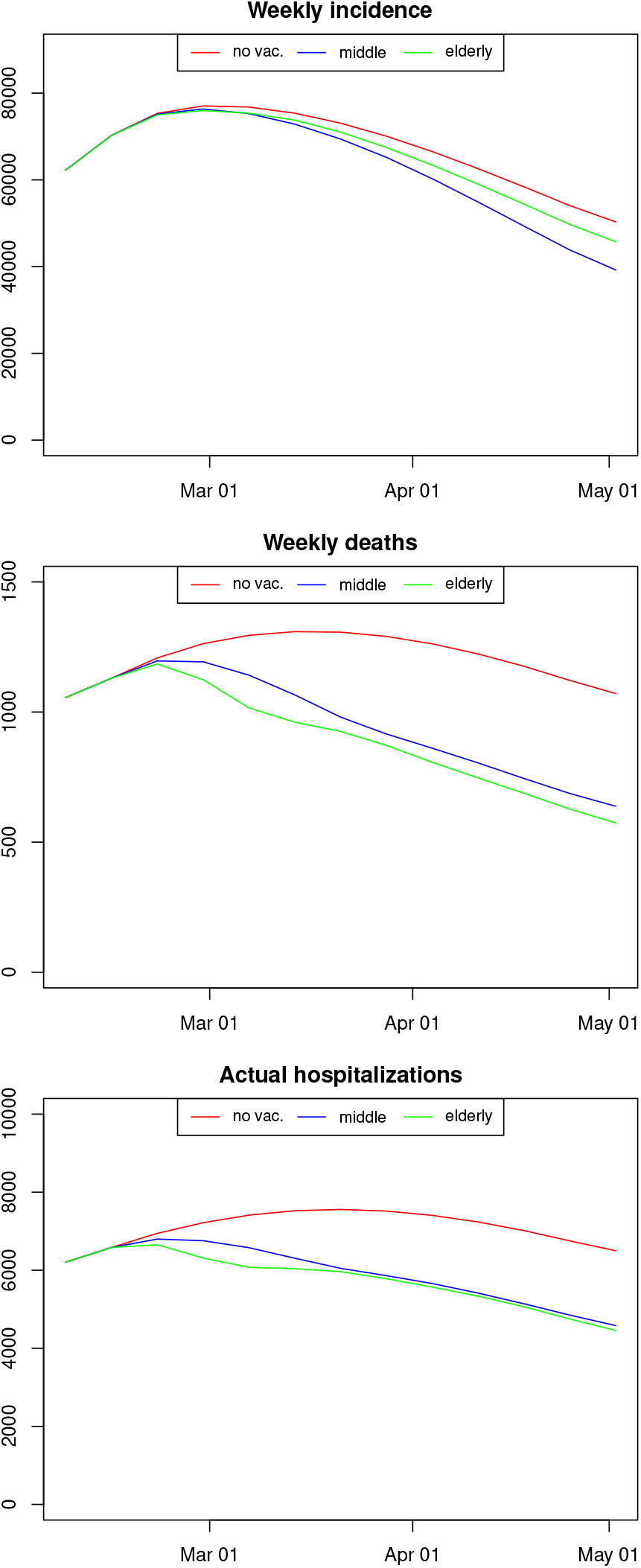
Outcomes of Vaccination Scenarios

## 11. Conclusion

We presented a stochastic epidemic model, formulated its basic properties and suggested way of its estimation, all demonstrated by a real-life examples. We demonstrated that our model has good predictive power. They are, however, many things to be done. Most importantly, a reliable automatic estimation procedure should be suggested, guaranteeing good fit of both means and variances. Second, regularity conditions which would ensure asymptotic properties should be formulated, yet it can be a difficult task. However, even as it is, it can be immediately used for statistically correct modeling of the present and perhaps also another epidemics.

## Data Availability

Data are available on github

https://github.com/cyberklezmer/seir/tree/ejor

## Appendix: Parameters of the Czech COVID Model

Table 2 lists the majority of parameters, some of them being estimated, the rest being determined. The following Table lists values, which we surveyed from literature in order to compute selected parameters.

|  | Description | Source |
| --- | --- | --- |
| $m_E = 5.08$ | mean of incubation period | [11] |
| $\alpha = 0.179$ | probability of asymptomatic course | [17] |
| $m_a = 4$ | expected duration of presymptomatic period | [18] |
| $m_i = 8$ | expected duration of infectiousness | [26] |

The parameters, not included in Table 2 are *θ*_*t*_, which is defined by 5, *π*_*t*_, which we define later, and ϖ _*t*_ which is defined by (8) but with owing explanation how *p*_*t*_ is computed.

The quantity *π*_*t*_ is set so that the overall detection probability, which we estimate by *δq*_*t*_ where *q*_*t*_ is discussed in the main text and *δ* is an estimated parameter. If we assume that, over time, the states of an individual follows a *continuous-time* Markov chain with transition matrix given by the parameters listed in Figure 2, then the probability of detection (excluding the cases when an undetected arrives to hospital or dies) is

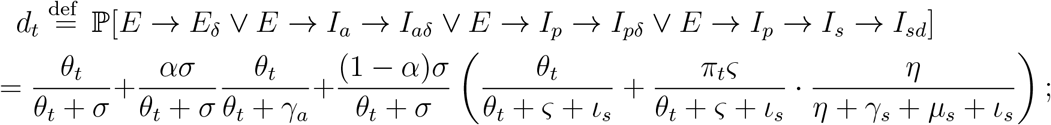

by putting *d*_*t*_ = *δq*_*t*_, we get *π*_*t*_ (trimming when *π*_1_ falls outside [0, 1]).

To evaluate the level of personal protection *p*_*t*_ is difficult task, as the study [20] monitors observance of several protective measures. However, if we assume that the *i*-th measure reduces the probability of infection by *λ*_*i*_, we get that, denoting 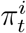 the average observance of the *i*-th measure among the respondents at *t*, that the average reduction brought by the the measure will be 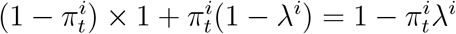. This gives total reduction

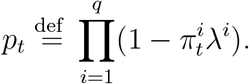

where *q* is the number of measures. Unfortunately, *λ*_*i*_ are unknown and their estimation would bring a serious danger of over-fitting and/or co-linearity (series 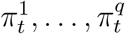. are almost perfectly corelated). To overcome this difficulty, we applied factor analysis to 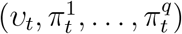 on the respondent level, treating the responses in different times as separate observations. As a result, we extracted two following main factors:

| $f$ | $g$ | Value |
| --- | --- | --- |
| 0.563 | 0.352 | Avoiding caughing people (yes or no) |
| 0.315 | 0.669 | Avoiding crowded places |
| 0.269 | 0.454 | Wearing a mask or a respirator |
| 0.394 | 0.610 | Restricting physical contact with people |
| 0.539 | 0.095 | Using desinfection |
| 0.526 | 0.295 | Avoiding people being in contact with an infected |
| 0.081 | 0.734 | Avoiding public transport |
| 0.417 | 0.100 | Taking vitamines |
| 0.640 | 0.201 | Avoiding touching nose and eyes |
| 0.565 | 0.261 | Extra hygiene |
| 0.661 | 0.121 | Washing hands after coughing |
| 0.670 | -0.244 | Washing hands after using public transport. |
| 0.084 | -0.517 | $w_t$ |

It can be seen that, while the first factor speaks more about contacts, the second one concerns personal protection, lacking connection with *υ*. Being interested in the protection, we approximate

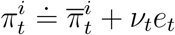

where 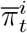 is the average of 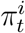 over time and respondents, *ν*_*t*_ is a constant and *e*_*t*_ is average of the second factor over respondents at *t*. Having that. we could approximate

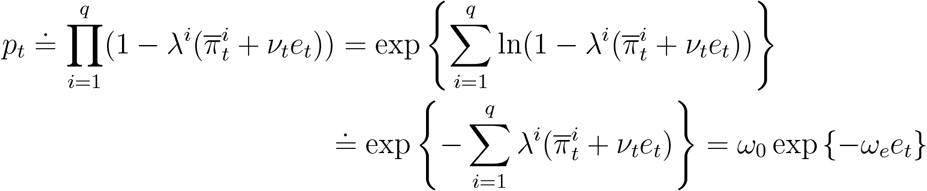

where *ω*_0_, *ω*_*e*_ ≥0. Similarly we can evaluate (assumed) reduction caused by the average reported fear *f*_*t*_ of infection, giving (7).

As for the adjustment *g*_*t*_ for the new virus variants, we assume *g*_*t*_ = 1 up to November 20, 2020, *g*_*t*_ = *ν*, where *ν* is an (estimated) parameter, since January 1, 2021, and that *g*_*t*_ is linearly interpolated between the dates. Yet the dates do not correspond to the common knowledge about mixing of the British variant, they come from a regression estimate of the reproduction number by *υ*.

Finally, we list the population of individual cohorts together with the “mixing matrix” *Q* (Table 3) and the definition of transformation matrix *F* (Figure 6).

**Figure 6:**
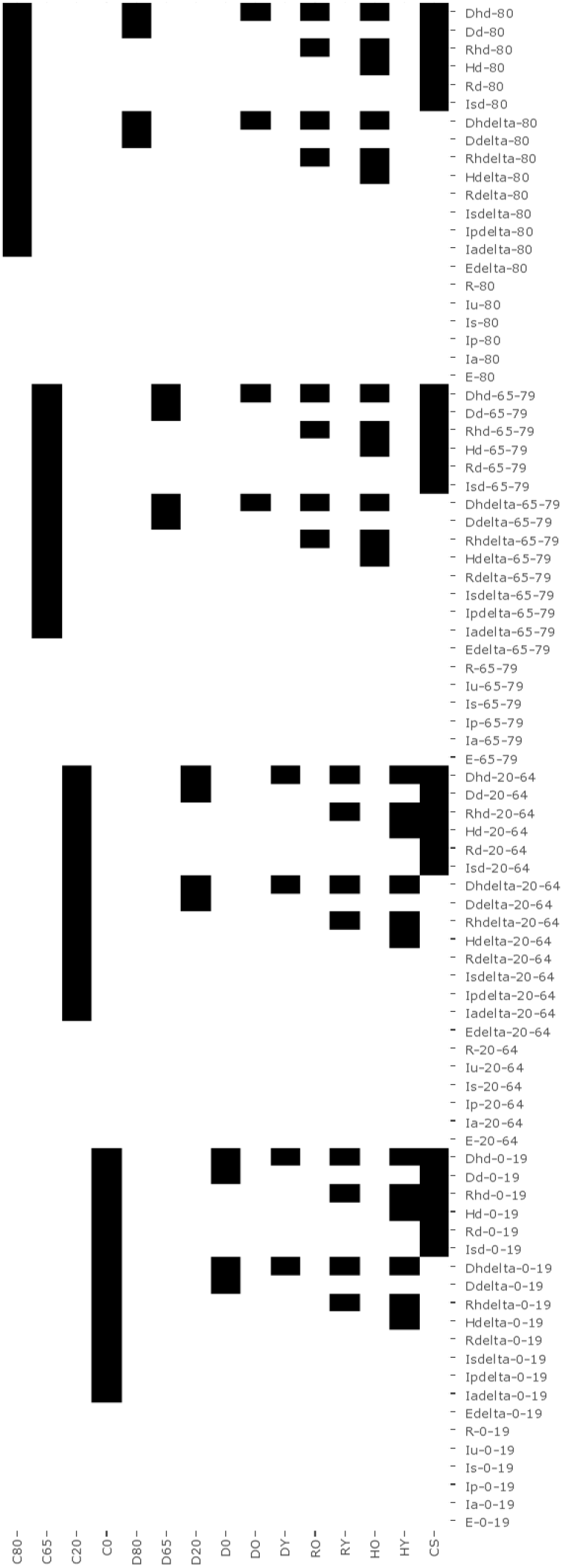
Definition of Transformation Matrix *F*

## Footnotes

7 It should be, however, noted that we do not prove asymptotic properties of the estimators.

8 In particular, *N*_*t +1,i*_ = CPo(*k* ln(1+*θ*); Log(p)), where Log is the Logarithmic distribution ([28]). Thus, 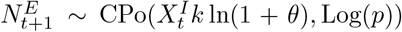 so we may put 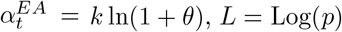.

